# Antibody persistency and trend post-SARS-CoV-2 infection at eight months

**DOI:** 10.1101/2020.11.21.20236117

**Authors:** Puya Dehgani-Mobaraki, Asiya Kamber Zaidi, Annamaria Porreca, Alessandro Floridi, Emanuela Floridi

## Abstract

An improved understanding of the immunity offered by the antibodies developed against SARS-CoV-2 is critical for the development of diagnostic tests and vaccines. Our study aimed at the longitudinal analysis of antibody presence, persistence and its trend over a period of eight months in a group of COVID-19 recovered patients who tested positive by real-time quantitative PCR for SARS-CoV-2 in the period between the 18th and 30th of March, 2020. The subjects were divided into two groups based on disease severity: mild and moderately-severe. The MAGLUMI 2019-nCoV lgM/lgG chemiluminescent analytical system (CLIA) assay was used to analyse the antibody titres. Robust IgG antibody persistency was demonstrated in 76.7 % of the subjects (23 out of 30) at eight months post-infection. The results of this study highlight an important point in terms of the association between humoral immune response and disease severity. Patients who might have experienced a relatively moderate-severe infection may develop a robust immunity that could persist for a longer duration.

## Introduction

Immunity offered by the antibodies developed in recovered COVID-19 patients is a matter of ongoing debate and encompasses genuine concern for the future. Recent studies and the media have focused their attention on the role of vaccines offering immunity and persistence of humoral response ranging from four months [1] to more than six months [2] post-infection.

## Methods

A monocentric pilot longitudinal observational study was conducted on 114 subjects based in the Umbria region in Italy, who had tested positive by real-time quantitative PCR for SARS-CoV-2 in the period between 18-30 March 2020. The demographic characteristics, blood groups, associated co-morbidities, clinical features, treatment undertaken and dates pertaining to symptom onset and swab collections were recorded. Sequential serum samples were collected over a period of eight months for 30 out of 114 subjects who attended all follow-up visits. The aim of this study was to investigate for presence, persistence and trend of IgM and IgG developed against SARS-CoV-2 over time. The subjects were divided into two groups based on disease severity: mild and moderately-severe. [3] The MAGLUMI® 2019-nCoV lgM/lgG chemiluminescent analytical system (CLIA) assay was used to analyse the antibody titres. Results were reported as measured chemiluminescence values divided by the cut off (absorbance/cut-off, S/CO): S/CO>1 was defined as positive and S/CO≤1 as negative. [4] We treated time as a factor and defined five different time points: first blood sample was collected in the month of May (T0) and then, one month (T1), three months (T2), five months (T3),six months (T4) after T0.

## Results

The descriptive statistics for the main characteristics of the study group was expressed as Median, [1_st_-3_rd_] quartile for continuous variables and as absolute frequency (column percentage) for the categorical variables. [Table 1A] Chi squared test was used to measure the association between disease severity and the categorical variables while the Mann U Whitney test was used to assess the differences between the disease severity for the continuous variables. Also, in order to analyse if the differences between disease severity at each time point were significant, the Mann U Whitney test was used. The Friedman test was applied to look for statistically significant differences over time between the two severity groups for IgM and IgG. [Table 1B] The normal distribution of the data was tested by Shapiro Wilks. Robust IgG antibody persistency was demonstrated in 76.7% of the subjects (23 out of 30) at eight months post infection. [FIGURE 1A]. The positivity cut-off for CLIA was set at 1.01 and the median titre trends were plotted for IgM and IgG, for both severity groups. [FIGURE 1B] For the mild group, the IgM titre trend stayed below the set cut off throughout time, but changed in a statistically significant way. The IgG titre trend dipped at T2 only to return to an almost linear trend at T3 but did not change in a statistically significant way. Similar findings were observed in previous studies. [5] For the moderately-severe group, the titre trend for both IgM and IgG changed in a statistically significant way throughout time with IgM below and IgG above the set cut-off. The IgG titre trend in this group dipped at T1 and peaked at T2. This dip experienced for both mild and moderately severe groups for IgG might be due to contraction of the immune response and may not indicate waning of immunity. [6]

**Table 1.** Descriptive statistics for the main characteristics of the study group was expressed as Median, [1st-3rd] quartile for continuous variables (A) and Friedman test for repeated measurements only for variables with a p-value <0.05 reporting the time pairs for which the test was statistically significant. PWC= pairwise comparisons.

| Variable | Mild | Moderately-Severe | p-value |
| --- | --- | --- | --- |
|  | N=17 | N=13 |  |
| Age (years) | 39.0<br>[30.0;59.0] | 55.0 [38.0;57.0] | 0.645 |
| SEX: |  |  | 0.708 |
| F | 10<br>(58.8%) | 9 (69.2%) |  |
| M | 7 (41.2%) | 4 (30.8%) |  |
| Blood Group: |  |  | 0.360 |
| Other | 15<br>(88.2%) | 9 (69.2%) |  |
| Zero | 2 (11.8%) | 4 (30.8%) |  |
| Healthcare worker: |  |  | 0.599 |
| NO | 9 (52.9%) | 9 (69.2%) |  |
| YES | 8 (47.1%) | 4 (30.8%) |  |

(B)
| Mild |  |  |  |  |  |  |  |
| --- | --- | --- | --- | --- | --- | --- | --- |
| Variable | T0 | T1 | T2 | T3 | T4 | p-value | PWC |
| IgM | 0.50<br>[0.46;0.52] | 0.61<br>[0.52;0.64] | 0.36<br>[0.18;0.53] | 0.23<br>[0.17;0.33] | 0.21<br>[0.18;0.35] | <0.001 | T0≠T2<br>T0≠T3<br>T0≠T4<br>T1≠T2<br>T1≠T3<br>T1≠T4 |
| IgG | 1.84<br>[0.68;3.54] | 1.81<br>[0.56;2.90] | 1.70<br>[0.88;5.02] | 3.05<br>[0.50;3.80] | 3.13<br>[1.05;3.66] | 0.205 |  |
| Moderate-Severe |  |  |  |  |  |  |  |
| Variable | T0 | T1 | T2 | T3 | T4 | p-value | PWC |
| IgM | 0.64<br>[0.64;1.07] | 0.70<br>[0.66;1.44] | 0.66<br>[0.24;0.84] | 0.59<br>[0.31;0.82] | 0.54<br>[0.25;0.81] | <0.001 | T0≠T2<br>T0≠T3<br>T0≠T4<br>T1≠T3<br>T1≠T4<br>T3≠T4 |
| IgG | 2.37<br>[1.14;20.7] | 1.48<br>[0.83;20.6] | 4.10<br>[2.38;16.1] | 2.25<br>[1.04;10.2] | 1.89<br>[0.83;8.53] | 0.013 | T0≠T4<br>T2≠T3<br>T2≠T4<br>T3≠T4 |

| Variable | Mild | Moderately-Severe | p-value |
| --- | --- | --- | --- |
|  | N=17 | N=13 |  |
| Age (years) | 39.0<br>[30.0;59.0] | 55.0 [38.0;57.0] | 0.645 |
| SEX: |  |  | 0.708 |
| F | 10 (58.8%) | 9 (69.2%) |  |
| M | 7 (41.2%) | 4 (30.8%) |  |
| Blood Group: |  |  | 0.360 |
| Other | 15 (88.2%) | 9 (69.2%) |  |
| Zero | 2 (11.8%) | 4 (30.8%) |  |
| Healthcare worker: |  |  | 0.599 |
| NO | 9 (52.9%) | 9 (69.2%) |  |
| YES | 8 (47.1%) | 4 (30.8%) |  |

**Figure 1.**
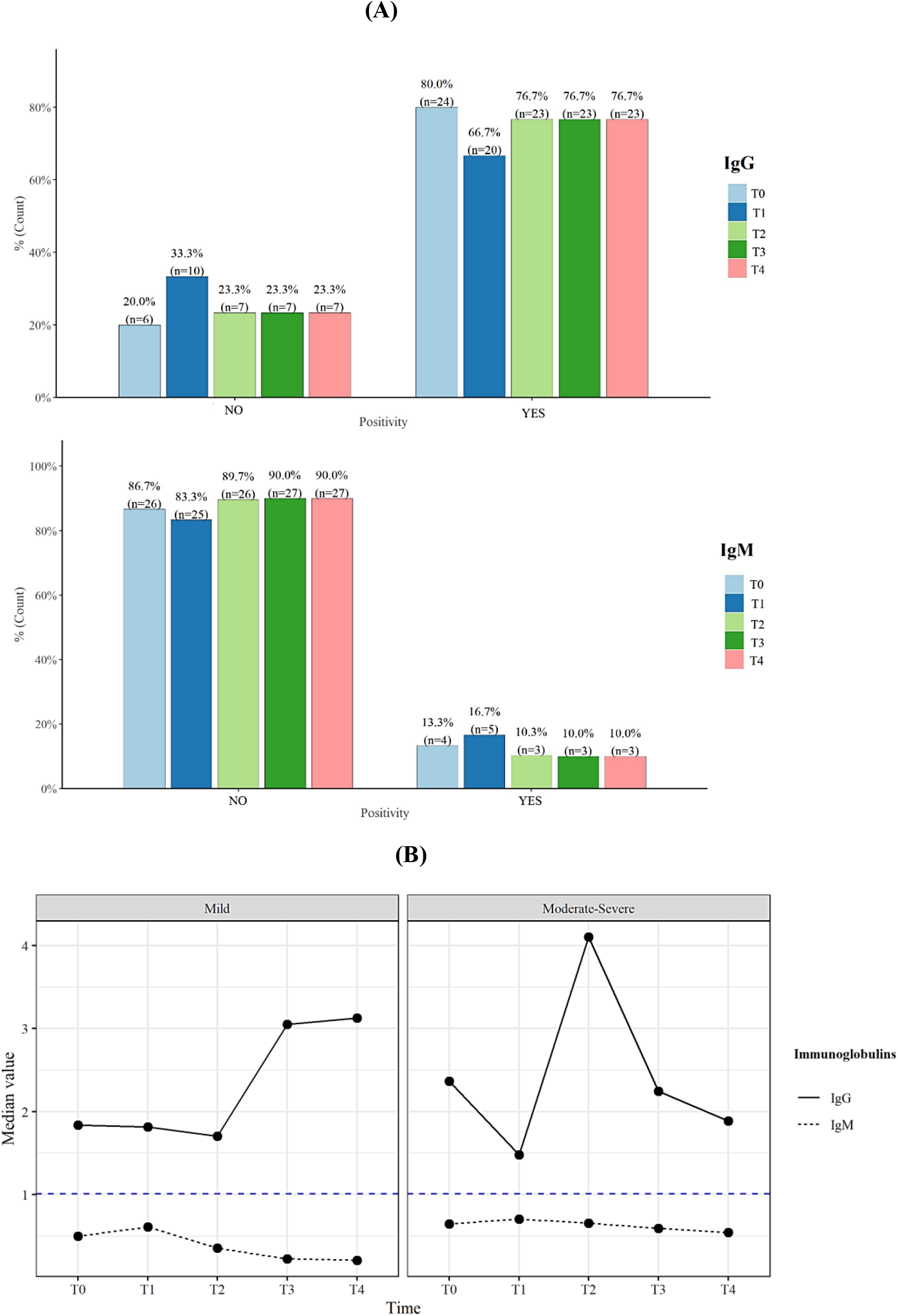
Bar plot showing IgM and IgG positivity for all subjects of the study group (N=30) (CLIA cut-off >1.00) in (A) and the Median trends for IgM and IgG for the two disease severity groups: mild and moderately severe. The dashed horizontal line shows the threshold (CLIA positivity Cut-off) for a sample being considered positive for antibodies in (B).

## Discussion

This study highlights an important point in terms of association between humoral immune response and disease severity. Patients who might have experienced a relatively severe infection may develop a robust immunity that could persist for a longer duration.

## Data Availability

Available on request

## Notes

### Competing Interest Statement

The authors have declared no competing interest.

### Clinical Trial

Observational study.

### Funding Statement

There was no funding source for this study.

### Author Declarations

This study was approved by the Research ethics committee of Association Naso Sano [Document number: ANS-2020/001].

